# Target Precision Shapes Trunk Compensation After Stroke: Neuromechanical Consequences for Upper Limb Rehabilitation

**DOI:** 10.64898/2026.10.01.26364468

**Authors:** Germain Faity, Denis Mottet, Jérôme Froger, Marion Delorme

**Author notes:** Correspondence: Germain Faity; ENS Rennes, Campus de Ker lann, 11 Av. Robert Schuman, 35170 Bruz, France.

## Abstract

**Background:** Paretic-arm nonuse and trunk compensation after stroke are often interpreted as impairment or learned behavior but may also reflect task-dependent optimization. We previously showed that gravitational shoulder effort and force reserve shape these behaviors. Here, we tested whether endpoint precision is an additional constraint.

**Methods:** Twenty-two individuals with stroke and 22 matched controls performed a seated Proximal Arm Nonuse reaching task toward small (2 cm) and large (12 cm) targets. Stroke participants were tested at normal arm weight and with the paretic arm mechanically lightened; controls were tested at normal arm weight and while holding a dumbbell corresponding to 85% of maximal voluntary shoulder force.

**Results:** In stroke participants, trunk compensation was 15% lower for the large target than for the small target, independently of arm weight condition (F(1,13)=8.166, p=.013, partial η²=.39). Target size did not significantly affect trunk compensation in controls or anterior deltoid activation in either group. In stroke participants, movement time was 24% longer for the small target than for the large target (F(1,13)=8.533, p=.012, partial η²=.40). In controls, the small target increased movement time only when the arm was weighted (F(1,18)=14.9, p=.001, partial η²=.45), not at normal arm weight (F(1,18)=0.158, p=.695, partial η²=.01).

**Conclusions:** Endpoint precision altered reaching after stroke partly independently of gravitational shoulder effort. Trunk compensation after stroke was therefore modulated by task demands rather than fixed by impairment, a pattern consistent with, although not a direct demonstration of, an optimal-control account. Rehabilitation should evaluate compensation by its functional role and the cost of suppressing it, not only by its deviation from typical kinematics, and should treat target size as an adjustable parameter of the task.

Clinical Trial: NCT04747587. https://clinicaltrials.gov/study/NCT04747587

## INTRODUCTION

Upper-limb rehabilitation after stroke is often organized around a distinction between recovery and compensation. Recovery denotes restitution of more typical movement capacity, whereas compensation denotes alternative neural or biomechanical strategies used to accomplish a task despite persistent impairment^1–3^. This distinction is useful, but it becomes clinically misleading when all compensatory movements are treated as equivalent. A forward trunk displacement during reaching can mask residual arm impairment during an impairment-oriented assessment. The same displacement can also help position the hand, avoid failure, or reduce the cost of a meaningful action.

Recent kinematic work has sharpened this issue by showing that well-recovered individuals after stroke may display near-normal distal endpoint kinematics while retaining altered proximal interjoint and intrajoint coordination^4^. Such findings indicate that successful endpoint performance can coexist with persistent proximal adjustments. They also raise a mechanistic and clinical question: should these adjustments be interpreted as residual deficits to be corrected, or can they sometimes contribute to successful reaching under the patient’s current neuromechanical constraints? This distinction is critical because the same proximal adjustment may be maladaptive in one patient and advantageous in another.

Paretic-arm nonuse has the same ambiguity. The learned nonuse framework proposes that repeated unsuccessful or aversive attempts to use the paretic limb can condition avoidance of that limb even when some capacity remains^5–7^. This hypothesis has shaped major therapeutic approaches and is consistent with use-dependent recovery principles after stroke^8–12^. However, arm use is not determined by capacity alone. Energetic cost and arm efficiency influence arm choice and real-world paretic-arm use^13,14^. Thus, beyond whether the paretic arm can accomplish a task, a complementary clinical question is whether using it is the most efficient solution under the current task constraints.

In a recent study using the same seated reaching paradigm, reducing the weight of the paretic arm decreased nonuse and trunk compensation after stroke. Conversely, increasing arm weight in controls induced trunk compensation of similar magnitude^15^. Reaching with spontaneous trunk compensation also reduced perceived exertion and preserved a shoulder force reserve^15^. These findings align with experimental and modeling work showing that, when a load becomes large relative to available shoulder torque, trunk recruitment can redistribute work to avoid excessive shoulder activation^16,17^.

These observations do not imply that nonuse is desirable, nor that compensation should be encouraged indiscriminately. They imply that poststroke reaching should be analyzed as a problem of action selection under constraints. The same patient may have enough motor capacity to reach a target with the paretic arm, yet still select a different coordination if the solution relying on the arm alone requires excessive relative activation or does not provide a sufficiently reliable way to meet the required endpoint tolerance. This distinction between capacity and efficiency is central to recent work showing that daily arm use after stroke is better predicted by arm efficiency than by capacity alone^14^.

Effort, however, is not the only constraint in reaching. In optimal feedback control, the nervous system is not assumed to reproduce a single normal kinematic pattern, but to select a control policy that satisfies the constraints of the task while minimizing an expected cost^18–20^. Depending on the task and the state of the neuromuscular system, this cost may include motor commands, movement time, endpoint variability, fatigue, discomfort, loss of force reserve, or risk of failure^17,21–23^. Endpoint precision is therefore not a secondary task feature: reducing target tolerance increases the cost of terminal error, so a coordination sufficient for a large target may be inadequate for a small one.

This precision constraint results in a trade-off between speed and accuracy: as target tolerance decreases, movement duration increases to preserve accuracy, a phenomenon quantified by Fitts’ law^24^. A plausible mechanism is that larger or faster motor commands tend to increase endpoint variability through signal-dependent noise^25^. More broadly, observed force and kinematic variability can also reflect feedback corrections and closed-loop control strategies, rather than motor noise alone^26^. Higher precision may therefore be achieved by slowing the movement, using corrective submovements, increasing limb impedance, or changing the coordination itself^27–31^. In a paretic limb, these accuracy-preserving options are constrained by weakness, abnormal coupling, altered activation, increased variability, and reduced smoothness^32–36^. Precision demand may therefore reveal compensatory strategies that cannot be explained by gravitational shoulder effort alone.

The present study tested this prediction using a target-size manipulation from a controlled seated reaching protocol. We asked whether smaller targets increase trunk compensation, movement time and perceived exertion in individuals with stroke and in controls. We hypothesized that high precision demand would increase trunk compensation and movement time after stroke, and that its persistence under arm lightening would indicate a precision-related control constraint distinct from gravitational shoulder effort control.

## METHODS

This controlled laboratory study examined how target size and arm-weight constraints shape paretic-arm use and trunk compensation during seated reaching after stroke. The arm-weight manipulation has been reported previously^15^. The present article focuses on the target-size manipulation from the same protocol.

### 1. Participants

Twenty-two adults with stroke and 22 age- and sex-matched controls were enrolled. Participants with stroke had a first supratentorial stroke, at least 1 month elapsed since stroke onset, and the ability to touch the opposite knee with the paretic arm while seated. Controls had no history of stroke and no upper-limb impairment, particularly at the shoulder. Exclusion criteria were inability to provide informed consent, cognitive difficulty preventing task understanding, or any other condition likely to affect task performance. Demographic, anthropometric, and clinical characteristics are reported in Table 1.

**Table 1.**
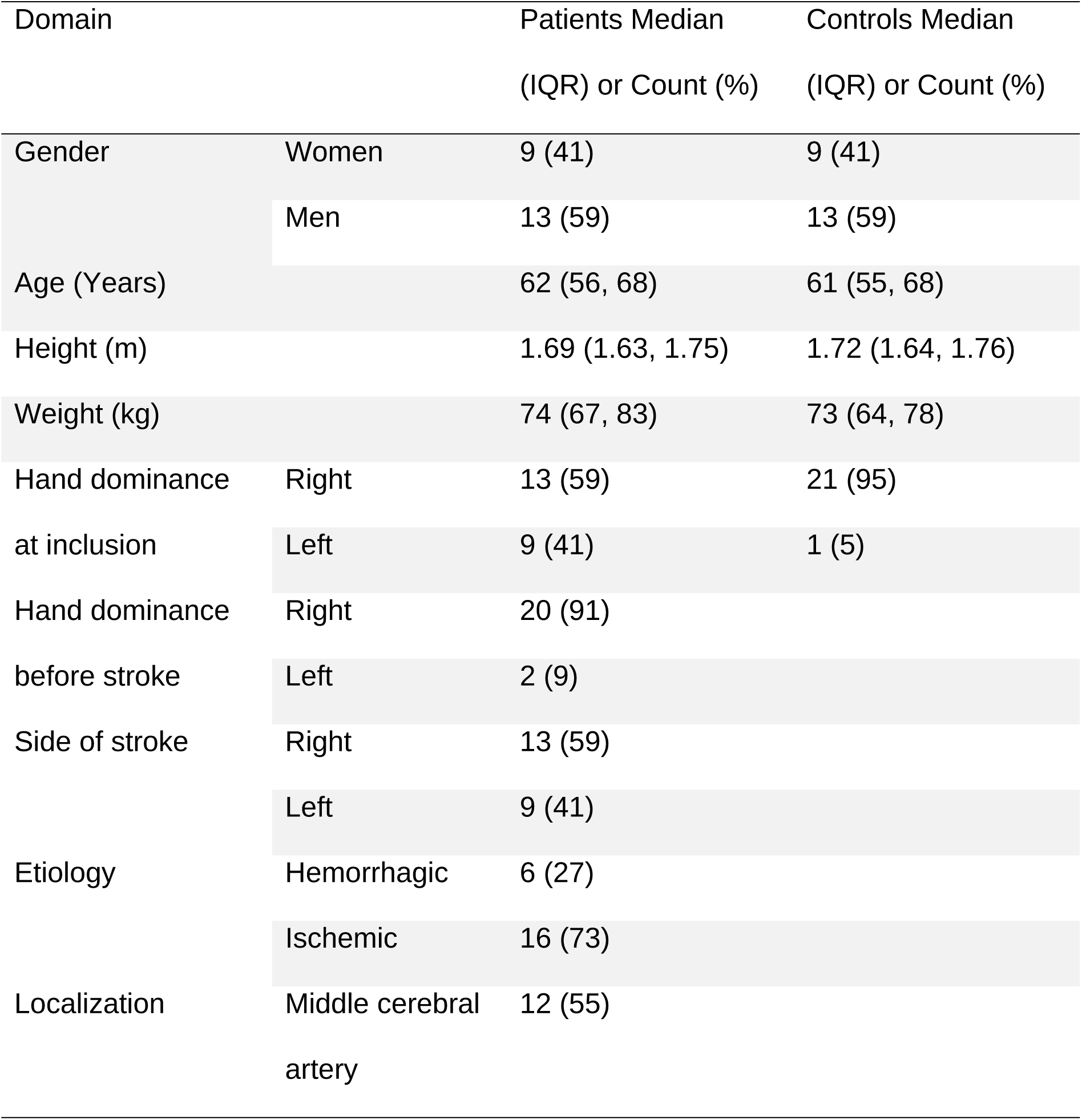
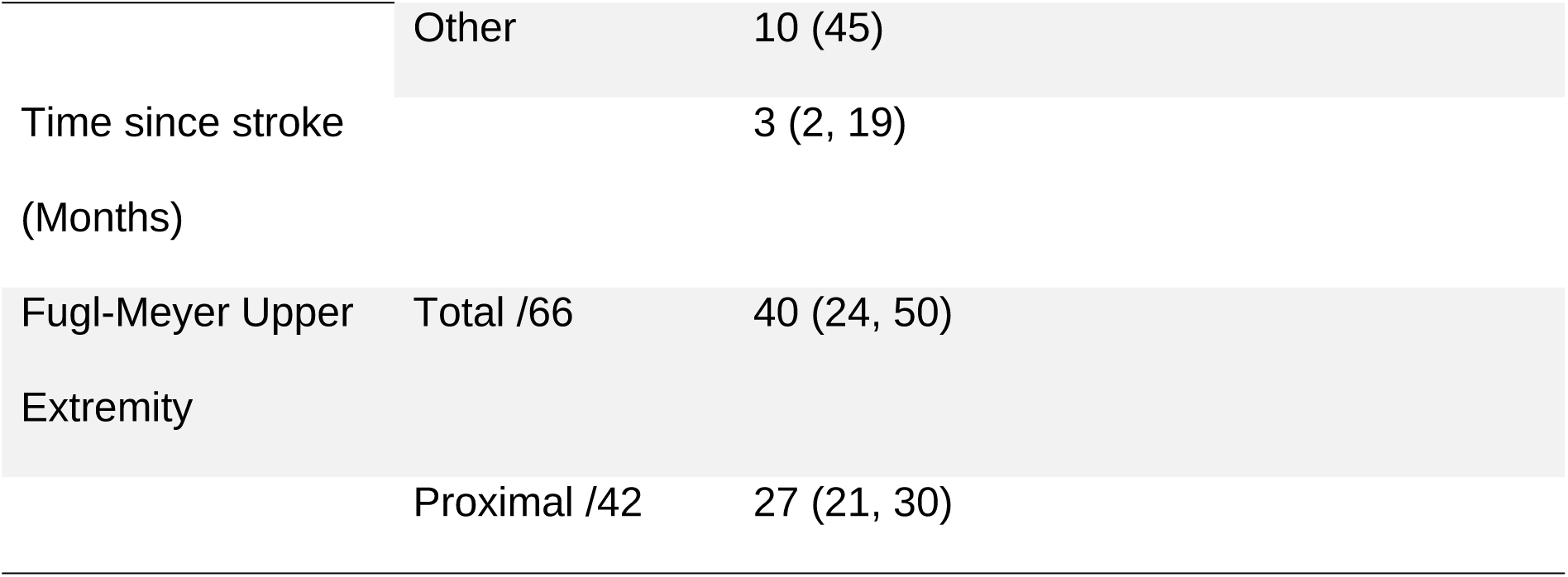
Demographic, anthropometric, and clinical characteristics of participants.

| Domain |  | Patients Median<br>(IQR) or Count (%) | Controls Median<br>(IQR) or Count (%) |
| --- | --- | --- | --- |
| Gender | Women | 9 (41) | 9 (41) |
|  | Men | 13 (59) | 13 (59) |
| Age (Years) |  | 62 (56, 68) | 61 (55, 68) |
| Height (m) |  | 1.69 (1.63, 1.75) | 1.72 (1.64, 1.76) |
| Weight (kg) |  | 74 (67, 83) | 73 (64, 78) |
| Hand dominance<br>at inclusion | Right | 13 (59) | 21 (95) |
|  | Left | 9 (41) | 1 (5) |
| Hand dominance<br>before stroke | Right | 20 (91) |  |
|  | Left | 2 (9) |  |
| Side of stroke | Right | 13 (59) |  |
|  | Left | 9 (41) |  |
| Etiology | Hemorrhagic | 6 (27) |  |
|  | Ischemic | 16 (73) |  |
| Localization | Middle cerebral<br>artery | 12 (55) |  |

|  |  |  |
| --- | --- | --- |
|  | Other | 10 (45) |
| Time since stroke<br>(Months) |  | 3 (2, 19) |
| Fugl-Meyer Upper<br>Extremity | Total /66 | 40 (24, 50) |
|  | Proximal /42 | 27 (21, 30) |

All participants provided written informed consent. The study was conducted in accordance with the 1964 Declaration of Helsinki and received approval from the Nimes Ethics Committee (N°ID-RCB: 2020-A02695-34, Clinical Trial: NCT04747587). The specific objective was not disclosed to participants to limit expectation bias.

### 2. Procedure

Participants performed the Proximal Arm NonUse (PANU) test^37^. The test includes 2 reaching conditions. In the Spontaneous Arm Use (SAU) condition, participants reached naturally toward a target positioned at individual arm length. In the Maximal Arm Use (MAU) condition, they were instructed to minimize trunk movement in order to maximize elbow and shoulder contribution. Trunk movement was minimized but not prevented, so that participants could still recruit the trunk when needed to reach the target. The difference between these conditions provides an operational estimate of nonmandatory trunk compensation and proximal arm nonuse. For each condition, participants performed 3 attempts, and the trial with the median trunk compensation was retained for analysis. A reach was considered successful when the hand touched the target. Unsuccessful reaches were rare, occurred on isolated attempts and were followed by a successful attempt, so task success was near ceiling in all conditions and was not analyzed as an outcome. During familiarization, all participants were able to touch the small target. Target distance corresponded to each participant’s arm length, measured from the medial axilla to the distal wrist crease.

Precision demand was manipulated using 2 spherical targets: a small 2-cm diameter target and a large 12-cm diameter target (Figure 1). The small target imposed a higher endpoint accuracy requirement, whereas the large target increased endpoint tolerance. Arm weight was also manipulated. All participants performed the task with their normal arm weight. Participants with stroke additionally performed the task with an arm-lightening device that counterbalanced the paretic arm. Controls additionally performed the task while holding a dumbbell corresponding to 85% of maximal voluntary shoulder force (MVF), a loading level used in our previous work and motivated by evidence that relative shoulder loading induces trunk recruitment when the arm becomes heavy relative to available force^15–17^. In the prior dumbbell paradigm, holding a negligible-weight dumbbell did not modify reaching coordination, supporting that the weighted condition primarily reflects relative shoulder loading rather than the mere act of holding an object^17^. Shoulder MVF was measured using the same isometric shoulder-flexion procedure as previously described^16^. Conditions were randomized and separated by rest periods to limit fatigue.

**Figure 1.**
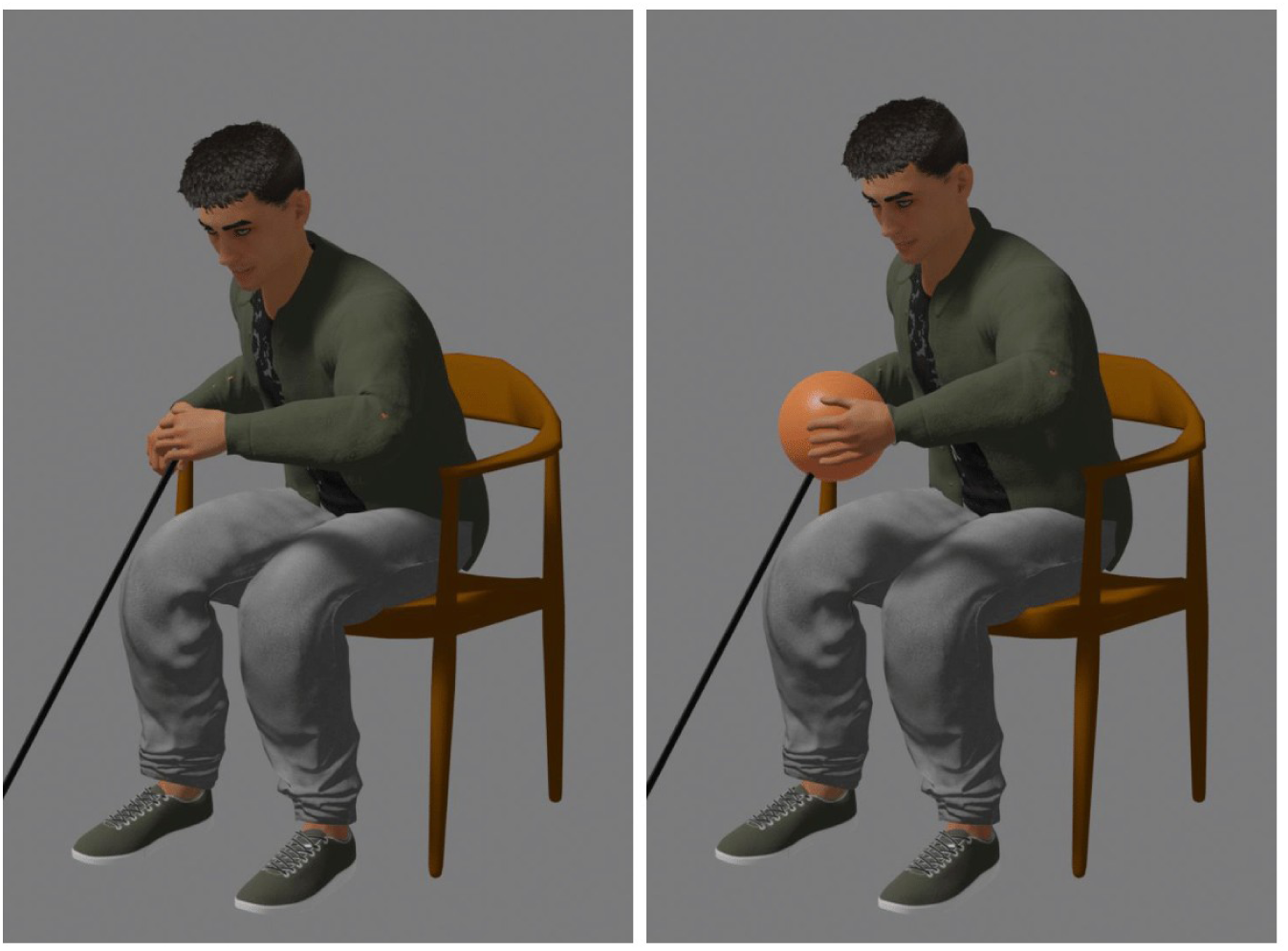
Experimental precision manipulation in the PANU reaching task. The same participant is shown reaching for the small target (left) and large target (right) in the spontaneous arm use condition with normal arm weight.

### 3. Experimental Setup

Movements were recorded at 30 Hz using the Kinect v2 markerless motion capture system (Microsoft, Redmond, WA), positioned 1.50 m in front of the participant. In this seated reaching paradigm, Kinect v2 provides sufficient validity and reliability for task-level outcomes such as shoulder displacement and movement time^38,39^. Surface EMG was recorded at 1258 Hz with a Delsys Trigno Wireless System. Electrodes were placed on the anterior deltoid, medial deltoid, biceps brachii, clavicular head of pectoralis major, and bilateral longissimus muscles. After each reaching task, participants rated perceived exertion on the Borg 6-to-20 scale^40^, which is known to accurately reflect actual effort in upper limb tasks^41^.

### 4. Data Processing

Data processing was carried out using Scilab 6.0.2.

Position time series were low-pass filtered at 2.5 Hz using a double-pass second-order Butterworth filter, a cutoff selected in the validation pipeline to retain most of the motion signal while reducing high-velocity tracking errors^39^. Movement onset was defined as the time at which Euclidean hand velocity became positive and remained positive until peak velocity. Movement end was defined as the time at which Euclidean distance between the hand and target reached its minimum^37^. Trunk compensation was quantified as the 3-dimensional Euclidean displacement of the shoulder between movement onset and movement end. In this seated reaching task, shoulder displacement provides a task-level estimate of trunk and shoulder-girdle recruitment during the reach^37^.

Electromyographic (EMG) signals were band-pass filtered between 10 and 850 Hz using a double-pass second-order Butterworth filter (80dB/dec). The root mean square (RMS) envelope was obtained with a 50-ms moving window applied to the absolute EMG signal. EMG was normalized to the corresponding MVC to estimate relative muscle activation. For each reach and each muscle, activation at the final posture was computed as the mean normalized EMG when the hand was within the final 5% of the reach length. The present manuscript focuses on anterior deltoid activation because it is a representative antigravity shoulder muscle in this reaching task but medial deltoid analyses led to the same interpretation.

### 5. Statistical analysis

Analyses were conducted in R 4.2.1 using the rstatix package (version 0.7.1). Normality was inspected with quantile-quantile plots. For each outcome, repeated-measures analyses tested target size (small versus large), arm-use condition (SAU versus MAU), and arm-weight context (stroke: normal versus lightened; controls: normal versus weighted). Significance was set at p<.05. Effect sizes are reported as partial η². Figures show means (± standard deviation). Full statistical results are reported in Supplemental Table S1.

## RESULTS

Trunk compensation was the primary outcome. Movement time and anterior deltoid activation were analyzed to test two explanations of the target-size effect: a speed-accuracy cost and a change in shoulder loading.

### 1. Target Size Modulated Trunk Compensation After Stroke

Target size affected trunk compensation in participants with stroke (Figure 2A). Shoulder displacement was 15% lower for the large target than for the small target across arm-weight contexts (target: F(1,13)=8.166, p=.013, partial η²=.39). This effect was not significantly modified by arm weight (arm weight x target: F(1,13)=0.084, p=.777, partial η²=.01), indicating that the target-size effect was present both when the paretic arm was tested at normal weight and when it was lightened. In controls, target size did not significantly affect trunk compensation (target: F(1,18)=0.057, p=.814, partial η²=.003), and the arm weight x target interaction was not significant (F(1,18)=0.681, p=.420, partial η²=.04).

**Figure 2.**
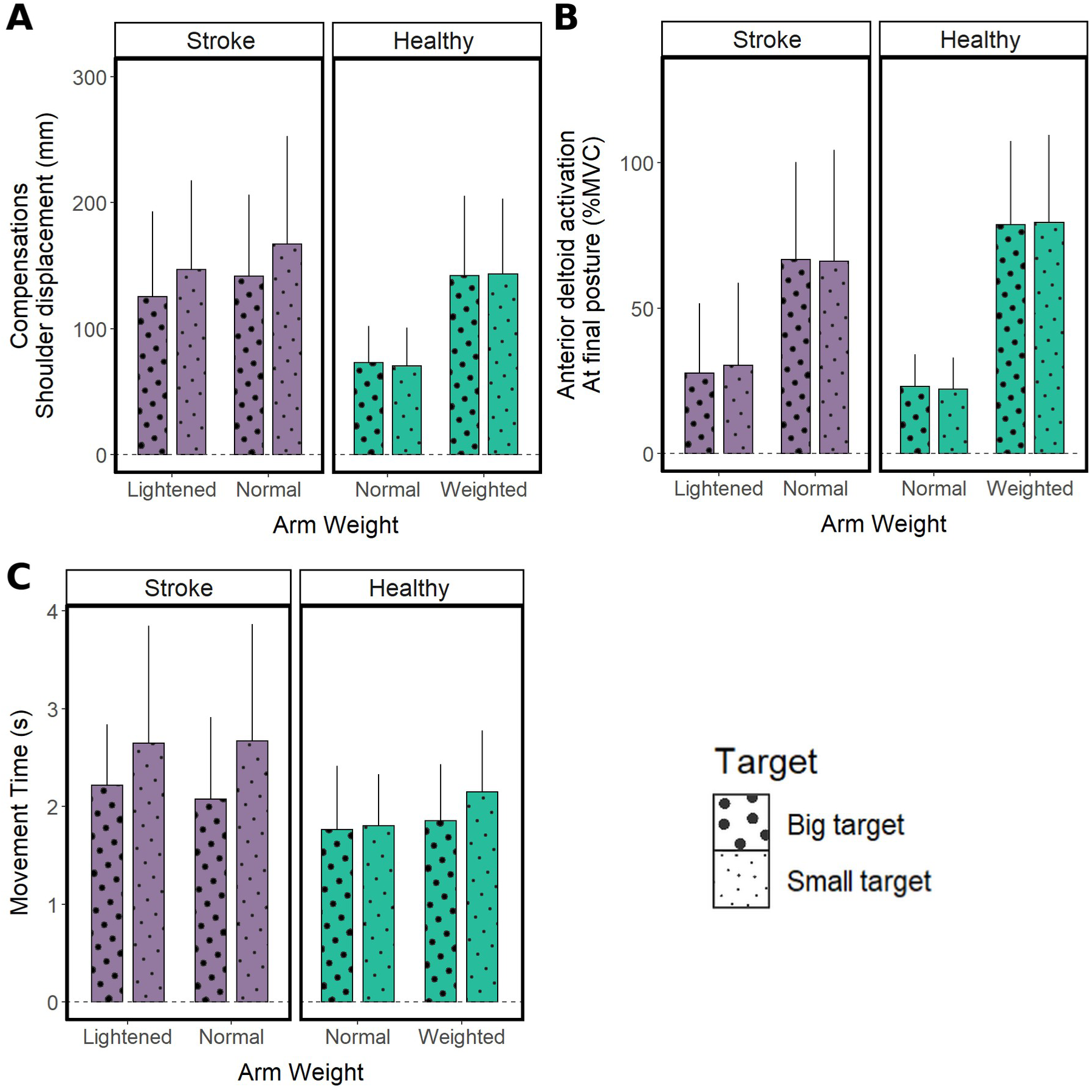
Influence of target size and arm-weight context on trunk compensation (A), anterior deltoid activation at the final posture (B), and movement time (C) in participants with stroke and controls. In participants with stroke, shoulder displacement was 15% lower for the large target and movement time was 24% longer for the small target, regardless of arm-weight context, whereas anterior deltoid activation was not affected. In controls, target size did not significantly affect trunk compensation or anterior deltoid activation, and increased movement time only when the arm was weighted. Data are presented as mean (± standard deviation).

Target-size effects did not depend on arm-use instruction in either group (all interactions involving target size and arm use, p≥.05; Supplemental Table S1). Thus, the previously described nonuse effect (SAU-MAU difference) was preserved across target sizes rather than modified by the precision manipulation.

### 2. Target Size Did Not Significantly Modify Anterior Deltoid Activation

The target-size effect on trunk compensation was not accompanied by a significant target-size effect on anterior deltoid activation (Figure 2B). Neither the main effect of target size (stroke: F(1,13)=0.149, p=.706, partial η²=.01; controls: F(1,18)=0.003, p=.960, partial η²<.001) nor any interaction involving target size (all p≥.05; Supplemental Table S1) was significant.

Medial deltoid analyses led to the same interpretation (data not shown).

### 3. Smaller Targets Increased Movement Time

Target size affected movement duration (Figure 2C). In participants with stroke, movement time was 24% longer for the small target than for the large target. The main effect of target size was significant (F(1,13)=8.533, p=.012, partial η²=.40), whereas the arm weight × target interaction was not significant (F(1,13)=0.316, p=.584, partial η²=.02). Participants with stroke therefore moved more slowly when endpoint tolerance was reduced, regardless of whether the paretic arm was tested at normal weight or mechanically lightened.

In controls, target size influenced movement duration only when reaching was made mechanically demanding by the added dumbbell. The main effect of target size was significant (F(1,18)=7.820, p=.012, partial η²=.30), whereas the arm weight × target interaction did not reach significance (F(1,18)=2.943, p=.103, partial η²=.14). With the arm weighted, movement time was 16% longer for the small target than for the large target (F(1,18)=14.9, p=.001, partial η²=.45). At normal arm weight, reaching duration was not significantly different between the small and large targets (F(1,18)=0.158, p=.695, partial η²=.01). Thus, in controls, the speed-accuracy cost emerged mainly when the arm was loaded, whereas in participants with stroke it was present even without added load.

Arm-use instruction did not significantly modify movement time or its dependence on target size in either group (all p≥.05; Supplemental Table S1).

Perceived exertion is reported in Supplemental Figure S1. It increased with the small target in controls (F(1,18)=9.500, p=.006, partial η²=.35), particularly when the arm was weighted, with a non-significant trend in participants with stroke (F(1,13)=4.304, p=.058, partial η²=.25). Perceived exertion therefore did not account for the stroke-specific increase in trunk compensation.

## DISCUSSION

Changing endpoint precision demand modified compensatory movements after stroke. In participants with stroke, trunk displacement was 15% lower for the large target than for the small target at both normal arm weight and with mechanical arm lightening. Smaller targets prolonged movement time by 24%, and the trunk effect was not accompanied by a detectable change in anterior deltoid activation. These findings extend our previous effort-based analysis by showing that poststroke compensation is shaped by at least two complementary constraints: the neuromechanical cost of moving a weakened arm and the control cost of producing a sufficiently accurate endpoint trajectory^15^.

### 1. Precision as an Optimal-Control Constraint

Beyond showing that smaller targets made the task harder, the key result is that endpoint precision changed the coordination selected to perform the reach. A fixed force-deficit account would predict similar trunk displacement across target sizes; a purely habitual learned-nonuse account would not predict sensitivity to current endpoint tolerance either. The target-size effect therefore indicates that poststroke trunk compensation retained task dependence. While this does not imply that all compensatory movements are useful, it cautions against viewing compensation as a mere deviation from typical kinematics.

These data are consistent with optimal feedback control, in which the motor system selects actions that achieve the goal with acceptable probability of success while limiting expected costs^18–22^. In a redundant musculoskeletal system, multiple trunk, shoulder, and elbow configurations can place the hand at the same location. The selected configuration should therefore depend on the relative weight assigned to effort, terminal error, and endpoint reliability^17^. When target tolerance decreases, the terminal-error term becomes more costly, and a coordination sufficient for a large target may no longer be reliable enough for a small one. The optimal coordination was not computed here, however, so these results are consistent with, rather than a direct demonstration of, optimization.

Signal-dependent variability offers one mechanism for this precision cost. Larger or faster motor commands increase endpoint variability, making slower movements advantageous when accuracy requirements are high^24,25^. Participants with stroke showed this temporal adjustment: movement time increased for the small target in both arm-weight contexts. Controls showed a comparable speed-accuracy adjustment only when the arm was made neuromechanically demanding by the added dumbbell. Thus, in the unweighted healthy system, the small target may have remained within the range of precision that could be achieved without a measurable change in global movement organization. In contrast, after stroke, the same reduction in target tolerance was imposed on a limb already characterized by weakness, abnormal interjoint coupling, altered activation, increased variability, and reduced smoothness^32–35^. Precision demand therefore acted on a system closer to its control limits.

Slowing movement is only one of the possible responses to an accuracy constraint. Accuracy can also be improved by increasing limb impedance through coactivation, which reduces endpoint sensitivity to neuromuscular noise and perturbations at the cost of greater metabolic demand and fatigue^28–31,42^. This possibility is considered below in relation to the anterior deltoid findings. When timing and impedance adjustments are not sufficient, or when they become too costly, the controller may instead reorganize the movement^27^. In this context, trunk recruitment can be interpreted as a change in coordination rather than as a simple failure of arm control. Indeed, shifting the trunk forward brings the shoulder closer to the target, thereby reducing the amount of endpoint displacement that must be generated by shoulder and elbow joints alone. The same hand location can then be reached with a different arm configuration, potentially lowering the relative demand on the paretic shoulder and increasing the stability of the final posture. Because trunk control generally benefits from bilateral innervation^1,3^, this strategy may help satisfy precision demands not only by reducing shoulder demand, but also by improving endpoint controllability. Consistent with this interpretation, failures were rare for both target sizes: participants with stroke preserved task success by adjusting movement time and coordination rather than by accepting more errors. The cost of the small target was therefore paid in time and trunk displacement, not in failure.

### 2. Why Arm Lightening Did Not Abolish the Precision Effect

In our previous work, arm lightening reduced effort-related compensation, whereas arm loading in controls induced stroke-like compensation^15^. Those findings identified gravitational shoulder effort and force reserve as major determinants of compensation. The present data add a boundary condition: reducing gravitational demand does not eliminate all compensatory adjustments. Precision remained able to modulate trunk recruitment even when the paretic arm was mechanically assisted.

This dissociation is consistent with the force-reserve model developed in healthy participants with induced shoulder strength deficits^17^. That model showed that trunk recruitment emerges as shoulder torque approaches a critical proportion of maximal capacity, because preserving a force reserve becomes more valuable than maintaining a reach without trunk compensation. The same model also predicted that shoulder weakness alone was insufficient to account for the full magnitude of poststroke compensation, implying additional constraints such as fine motor control, somatosensory reliability, spasticity, pain, or endpoint precision^17^. The present experiment directly tests one of these missing constraints. It shows that endpoint precision can modulate compensation after stroke even when the mechanical consequence of arm weight is reduced.

The anterior deltoid findings further constrain the interpretation. If the greater trunk displacement observed for the smaller target primarily reflected mechanical unloading of the shoulder, lower anterior deltoid activation would be expected in that condition. Instead, anterior deltoid activation was not detectably modified by target size. Preserved deltoid activation despite greater trunk recruitment is compatible with an accuracy-related stabilization strategy, potentially involving impedance modulation^28^ through augmented coactivation, consistent with the impedance-based account outlined above. This interpretation remains indirect, because the EMG montage primarily sampled agonist muscles and did not allow antagonist coactivation to be quantified. Future work combining agonist-antagonist EMG and endpoint-variability analyses will be needed to determine whether preserved deltoid activation reflects coactivation used to reduce variability or another component of the precision-related control policy.

### 3. Relation to Proximal Arm NonUse

The same framework applies to proximal arm nonuse. In the PANU task, the spontaneous arm-use condition allows participants to select their preferred coordination, whereas the maximal arm-use condition estimates the extent to which trunk contribution can be voluntarily reduced^37^. The persistence of the nonuse effect across target sizes indicates that precision demand did not generate proximal arm nonuse *de novo*. Rather, it modulated the expression of an already available coordination policy.

This distinction separates voluntary reducibility from functional value. A compensatory pattern may be nonmandatory, in the sense that it can be reduced when explicitly instructed, yet still provide a local advantage in the task being performed. Suppressing such a pattern may increase effort^15,17^, reduce endpoint precision, or increase failure risk. Conversely, a reducible compensation may be unnecessary when it no longer improves success, effort, or reliability. This duality may help explain why constraint-induced movement therapy is most feasible and effective in selected patients with sufficient residual function and engagement^6,43,44^.

In other words, trunk compensation may be the most efficient solution for the patient at a given time: it may minimize a composite cost^23^ combining effort and error under current impairments and task demands, without being the best option for long-term recovery. Habitual motor behavior and local minima may also limit exploration of more efficient solutions^45,46^. The clinical problem is therefore not whether compensation deviates from typical kinematics, but whether it still provides a measurable functional advantage under present constraints^15^. The target-size manipulation offers a way to probe this distinction by testing whether compensation remains sensitive to task demands.

### 4. Implications for Rehabilitation

Our perspective refines the interpretation of recent findings showing that endpoint kinematics may recover while proximal coordination remains altered after stroke^4^. Such a dissociation supports the idea that proximal adjustments may be relevant therapeutic targets, but it should not imply that they should be systematically suppressed. It may also indicate that endpoint performance is preserved through an alternative coordination strategy. The present results provide one way to test this distinction experimentally: if compensation varies with precision demand, load, assistance, or success criteria, it is more likely to reflect current task constraints; if it persists without measurable benefit for success, effort, or accuracy, it may represent a stronger candidate for compensation-reduction interventions.

In terms of clinical practice, these results also inform on the relative importance of task constraints. In the same patients, enlarging the target reduced trunk compensation by about 15%, a magnitude comparable to the 13% reduction obtained when the weight of the paretic arm was fully compensated^15^, and the two effects did not interact. Precision is therefore not a secondary detail compared with arm weight: in patients with mild-to-moderate impairment, both constraints weigh on compensation to a similar extent and appear to act additively. However, even with the large target and the lightened arm, trunk compensation after stroke remained above control values (Figure 2A), as previously observed with arm lightening^15^. Arm weight and precision therefore do not account for all poststroke compensation, and other determinants, such as impaired interjoint coordination or habitual behavior, likely contribute.

We propose that therapists consider target tolerance a dose parameter in upper-limb rehabilitation after stroke. Like load or number of repetitions, this dose can be quantified and graded: target width (W) and distance (D) define Fitts’ index of difficulty, ID = log_2_(2D/W)^24^, and the relation between ID and movement time can be measured in the paretic arm^35^, which makes it possible to monitor precision demand across sessions. Increasing this dose by reducing target size raises the cost of endpoint error. It may therefore alter movement time, perceived exertion, and the relative value of trunk recruitment. When the therapeutic objective is arm-specific practice with minimal trunk involvement, an excessive precision dose may shift the patient toward compensation, avoidance, or failure. Conversely, when the objective is successful functional performance under realistic constraints, some degree of compensation may be appropriate if it preserves success without excessive cost.

Accordingly, interventions aimed at reducing compensation remain relevant when compensation provides little gain in success, effort, or reliability. Trunk restraint, constraint-induced movement therapy, and task-specific practice may be appropriate in such cases^11,12,47–49^. This profile is consistent with learned nonuse, in which a familiar coordination persists despite improved capacity^15,45,46^. However, when compensation preserves task success, reduces effort, or improves endpoint control, suppressing it without modifying the underlying neuromechanical constraints may increase failure or reinforce costly movement. In this context, targeting shoulder force with resistance training, progressively reducing gravitational assistance, or using graded task design might be more appropriate initial targets, as suggested in previous work^15,17,50^. This interpretation is consistent with evidence that real-world arm use after stroke is better predicted by arm efficiency than by arm capacity alone^14^. Table 2 organizes these principles into a stepwise bedside sequence, from the question of whether compensation is mandatory to the choice of the constraint to target.

**Table 2.** Proposed sequence for interpreting and targeting trunk compensation during reaching after stroke. The sequence is derived from the present and previous findings^15,17^. It is hypothesis-generating and requires prospective validation before clinical use.

| Step | Clinical question and bedside test | Interpretation and suggested orientation |
| --- | --- | --- |
| **1. Is compensation mandatory?** | Ask the patient to reach a large target placed at arm length while keeping the trunk against the backrest, and compare with spontaneous reaching (as in the PANU test<sup>37</sup>). | Target not reached without the trunk: compensation is currently necessary. Prioritize capacity (shoulder strength, graded arm-weight support); avoid trunk restraint at this stage. |
| **2. What does suppressing it cost?** | If compensation is reducible, compare spontaneous and instructed reaching for success, movement time and perceived exertion (Borg scale). | Little or no cost: compensation brings no measurable benefit (profile consistent with learned nonuse). Compensation-reduction interventions are indicated (trunk restraint, constraint-induced movement therapy, task-specific training).<br>Clear cost (failure, slowing, higher exertion): compensation is currently useful; go to step 3 before restraining it. |
| **3. Which constraint makes it useful?** | Repeat spontaneous reaching with arm-weight support (sliding on a table, sling or spring-based support) and with a larger target. | Decreases with support: effort-related. Target shoulder strength and progressively reduce assistance<sup>50</sup>.<br>Decreases with a larger target: precision-related. Start practice without trunk compensation using large targets and reduce target size progressively.<br>Both effects may coexist. Other constraints, such as pain or sensory impairment, may also contribute. |
| **4. How to progress?** | Change one constraint at a time (load, assistance, target size) and repeat steps 1–3 periodically. | Increase precision or load only when success is maintained with limited trunk displacement. Interpret a return of compensation as a sign that the task has become too costly, not only as a fault to be corrected. |

A practical implication is that clinicians should manipulate target size deliberately rather than treating it as a neutral task parameter. Larger targets may facilitate paretic-arm contribution while limiting the need for trunk recruitment. Smaller targets may be useful to challenge endpoint stabilization, provided that success remains attainable and compensation is interpreted rather than simply penalized. The same patient may therefore require different target tolerances depending on whether the session aims to promote reaching without trunk compensation, functional task success, shoulder force- reserve training, endpoint precision, or transfer to daily activities. Precision, load, assistance, and success criteria should be adjusted jointly, because each of these parameters changes the cost structure of the task. For example, in a patient whose trunk displacement decreases with a larger target, practice without trunk compensation could begin with large targets and move to smaller ones only once reaching is performed with little trunk displacement and without loss of success.

For future clinical research, these findings support evaluating compensation according to the function it serves. A reaching movement with large trunk displacement does not have the same meaning across target sizes, loads, assistance levels, and success criteria. Future intervention studies could stratify patients according to whether trunk recruitment improves success, reduces effort, or improves accuracy under specific task constraints, as suggested before^15^. Patients who derive little measurable benefit from compensation might be the best candidates for compensation-reduction strategies. Patients whose compensation preserves precision or reduces effort may require changes in strength, sensory reliability, task difficulty, or assistance before compensation can be reduced without increasing risk of failure or effort. The sequence proposed in Table 2 should be evaluated in such trials before being considered as a clinical guideline.

### 5. Limitations

This study has several limitations. First, endpoint precision was manipulated using only two target sizes. A broader range of target tolerances would allow formal estimation of speed-accuracy functions and would make it possible to determine whether precision- compensation relationships differ across patients. Second, EMG interpretation was based on amplitude measures from a limited set of muscles. The absence of a target-size effect on anterior deltoid activation therefore does not exclude antagonist coactivation. Future studies should combine agonist-antagonist EMG with impedance-related measures to test these mechanisms directly. Third, Kinect v2 is known to be appropriate for the task-level outcomes analyzed here, particularly shoulder displacement and movement time^38,39^, but it does not provide the spatial and temporal resolution required to characterize subtle changes in joint coordination, movement smoothness, endpoint error, trial-by-trial variability, or corrective submovements. Gold-standard motion capture combined with direct endpoint-accuracy measures will be needed to determine whether trunk recruitment actually improves precision, reduces variability, or accompanies another accuracy-preserving strategy. Finally, the sample was heterogeneous but mostly centered on mild-to-moderate upper-limb impairment. Patients with severe paresis, major sensory loss, neglect, pain, spasticity, or cognitive impairment may operate under different cost structures and may show different relationships between precision demand and compensation.

## CONCLUSION

Endpoint precision changed compensatory behavior in seated reaching after stroke. Larger targets reduced trunk compensation, whereas smaller targets increased movement duration. Trunk compensation after stroke was thus modulated by task demands rather than fixed by impairment. These findings support a neuromechanical interpretation of proximal arm nonuse: trunk compensation depends on the task and constitutes an adaptive strategy that allows individuals to meet precision and effort demands when motor capacity is impaired. Rehabilitation should evaluate compensation by the function it serves and by the cost of removing it, not merely by deviation from typical kinematics. In practice, target tolerance should be adjusted deliberately, together with arm load and assistance, when the aim is to promote reaching without trunk compensation.

## ACKNOWLEDGMENTS

We would like to thank Simon Pla and Hugo Rodriguez for helping set up the experiment.

## DATA AVAILABILITY

The code and the datasets generated and analyzed during the current study are available in the Open Science Framework repository, https://osf.io/4mr7k/.

## AUTHOR CONTRIBUTIONS

Conceptualization, G.F., D.M., and J.F.; methodology, G.F., D.M., and J.F.; software, G.F. and D.M.; validation, G.F., D.M., J.F., and M.D.; formal analysis, G.F. and D.M.; investigation, G.F.; resources, D.M., J.F., and M.D.; data curation, G.F.; writing-original draft, G.F.; writing-review and editing, G.F., D.M., J.F., and M.D.; visualization, G.F. and D.M.; supervision, D.M. and J.F.; project administration, G.F., D.M., J.F., and M.D.; funding acquisition, G.F., D.M., and J.F. All authors have read and agreed to the published version of the manuscript.

## ABBREVIATIONS

PANU-: Proximal Arm NonUse
SAU-: Spontaneous Arm Use
MAU-: Maximal Arm Use
MVF-: Maximal Voluntary Force
EMG-: Electromyography
RPE-: Rate of Perceived Exertion

## REFERENCES

1. Cirstea MC, Levin MF. Compensatory strategies for reaching in stroke. Brain. 2000;123(5):940–953. doi:10.1093/brain/123.5.940

2. Levin MF, Kleim JA, Wolf SL. What Do Motor “Recovery” and “Compensation” Mean in Patients Following Stroke? Neurorehabil Neural Repair. 2009;23(4):313–319. doi:10.1177/1545968308328727

3. Jones TA. Motor compensation and its effects on neural reorganization after stroke. Nat Rev Neurosci. 2017;18(5):267–280. doi:10.1038/nrn.2017.26

4. Wolf S, Winter LV, Elangovan N, et al. Compensatory Proximal Adjustments Characterize Effective Reaching Movements After Stroke. Stroke. 2025;56(8):2245–2254. doi:10.1161/STROKEAHA.124.049336

5. Edward Taub. Somatosensory Deafferentation Research with Monkeys Implications for Rehabilitation. Behavioral Psychology in Rehabilitation Medicine: Clinical Applications. 1980;(ed. Ince, L. P.).

6. Taub E, Uswatte G, Elbert T. New treatments in neurorehabiliation founded on basic research. Nat Rev Neurosci. 2002;3(3):228–236. doi:10.1038/nrn754

7. Hirsch T, Barthel M, Aarts P, et al. A First Step Toward the Operationalization of the Learned Non-Use Phenomenon: A Delphi Study. Neurorehabil Neural Repair. 2021;35(5):383–392. doi:10.1177/1545968321999064

8. Ballester BR, Winstein C, Schweighofer N. Virtuous and Vicious Cycles of Arm Use and Function Post-stroke. Frontiers in Neurology. 2022;13. Accessed April 20, 2023. https://www.frontiersin.org/articles/10.3389/fneur.2022.804211

9. Han CE, Arbib MA, Schweighofer N. Stroke Rehabilitation Reaches a Threshold. PLOS Computational Biology. 2008;4(8):e1000133. doi:10.1371/journal.pcbi.1000133

10. Hidaka Y, Han CE, Wolf SL, Winstein CJ, Schweighofer N. Use It and Improve It or Lose It: Interactions between Arm Function and Use in Humans Post-stroke. Diedrichsen J, ed. PLoS Computational Biology. 2012;8(2):e1002343. doi:10.1371/journal.pcbi.1002343

11. Wolf SL, Winstein CJ, Miller JP, et al. Effect of constraint-induced movement therapy on upper extremity function 3 to 9 months after stroke: the EXCITE randomized clinical trial. JAMA. 2006;296(17):2095–2104. doi:10.1001/jama.296.17.2095

12. Corbetta D, Sirtori V, Castellini G, Moja L, Gatti R. Constraint-induced movement therapy for upper extremities in people with stroke. Cochrane Database Syst Rev. 2015;(10). doi:10.1002/14651858.CD004433.pub3

13. Nguyen H, Phan T, Shadmehr R, Lee SW. Choice of Arm Use in Stroke Survivors is Largely Driven by the Energetic Cost of the Movement. Neurorehabil Neural Repair. 2023;37(4):183–193. doi:10.1177/15459683231164788

14. Le Perf G, Faity G, Mottet D, Muthalib M, Laffont I, Bakhti K. Beyond Arm Capacity in Chronic Stroke: Evaluating Paretic Arm Non-Use Through Arm Efficiency—A Cross-Sectional Study. Neurorehabil Neural Repair. 2025;39(6):423–432. doi:10.1177/15459683241303691

15. Faity G, Mottet D, Froger J, Delorme M. Neuromechanics of Nonuse and Compensation: Implications for Rehabilitation After Stroke. Neurorehabil Neural Repair. 2026;40(8):615–630. doi:10.1177/15459683261445442

16. Faity G, Mottet D, Pla S, Froger J. The reserve of joint torque determines movement coordination. Sci Rep. 2021;11(1):23008. doi:10.1038/s41598-021-02338-4

17. Faity G, Barradas VR, Schweighofer N, Mottet D. Force reserve predicts compensation in reaching movement with induced shoulder strength deficit. J Neurophysiol. 2024;132(2):470–484. doi:10.1152/jn.00143.2024

18. Todorov E, Jordan MI. Optimal feedback control as a theory of motor coordination. Nature Neuroscience. 2002;5(11):1226–1235. doi:10.1038/nn963

19. Diedrichsen J, Shadmehr R, Ivry RB. The coordination of movement: optimal feedback control and beyond. Trends Cogn Sci. 2010;14(1):31–39. doi:10.1016/j.tics.2009.11.004

20. Scott SH. Optimal feedback control and the neural basis of volitional motor control. Nat Rev Neurosci. 2004;5(7):7. doi:10.1038/nrn1427

21. Rigoux L, Guigon E. A Model of Reward- and Effort-Based Optimal Decision Making and Motor Control. PLoS Comput Biol. 2012;8(10):e1002716. doi:10.1371/journal.pcbi.1002716

22. Shadmehr R, Huang HJ, Ahmed AA. A Representation of Effort in Decision-Making and Motor Control. Current Biology. 2016;26(14):1929–1934. doi:10.1016/j.cub.2016.05.065

23. Berret B, Chiovetto E, Nori F, Pozzo T. Evidence for Composite Cost Functions in Arm Movement Planning: An Inverse Optimal Control Approach. PLOS Computational Biology. 2011;7(10):e1002183. doi:10.1371/journal.pcbi.1002183

24. Fitts PM. The information capacity of the human motor system in controlling the amplitude of movement. Journal of Experimental Psychology. 1954;47(6):381–391. doi:10.1037/h0055392

25. Harris CM, Wolpert DM. Signal-dependent noise determines motor planning. Nature. 1998;394(6695):780. doi:10.1038/29528

26. Nagamori A, Laine CM, Loeb GE, Valero-Cuevas FJ. Force variability is mostly not motor noise: Theoretical implications for motor control. PLOS Computational Biology. 2021;17(3):e1008707. doi:10.1371/journal.pcbi.1008707

27. Tseng YW, Scholz JP, Schöner G, Hotchkiss L. Effect of accuracy constraint on joint coordination during pointing movements. Exp Brain Res. 2003;149(3):276–288. doi:10.1007/s00221-002-1357-5

28. Selen LPJ, van Dieën JH, Beek PJ. Impedance Modulation and Feedback Corrections in Tracking Targets of Variable Size and Frequency. Journal of Neurophysiology. 2006;96(5):2750–2759. doi:10.1152/jn.00552.2006

29. Tanis D, Calalo JA, Cashaback JGA, Kurtzer IL. Accuracy and effort costs together lead to temporal asynchrony of multiple motor commands. J Neurophysiol. Published online November 30, 2022:jn.00435.2022. doi:10.1152/jn.00435.2022

30. Burdet E, Osu R, Franklin DW, Milner TE, Kawato M. The central nervous system stabilizes unstable dynamics by learning optimal impedance. Nature. 2001;414(6862):446–449. doi:10.1038/35106566

31. Gribble PL, Mullin LI, Cothros N, Mattar A. Role of Cocontraction in Arm Movement Accuracy. Journal of Neurophysiology. 2003;89(5):2396–2405. doi:10.1152/jn.01020.2002

32. Dewald JPA, Pope PS, Given JD, Buchanan TS, Rymer WZ. Abnormal muscle coactivation patterns during isometric torque generation at the elbow and shoulder in hemiparetic subjects. Brain. 1995;118(2):495–510. doi:10.1093/brain/118.2.495

33. Beer RF, Dewald JPA, Rymer WZ. Deficits in the coordination of multijoint arm movements in patients with hemiparesis: evidence for disturbed control of limb dynamics. Experimental Brain Research. 2000;131(3):305–319. doi:10.1007/s002219900275

34. McCrea PH, Eng JJ, Hodgson AJ. Saturated Muscle Activation Contributes to Compensatory Reaching Strategies After Stroke. J Neurophysiol. 2005;94(5):2999–3008. doi:10.1152/jn.00732.2004

35. McCrea PH, Eng JJ. Consequences of increased neuromotor noise for reaching movements in persons with stroke. Exp Brain Res. 2005;162(1):70–77. doi:10.1007/s00221-004-2106-8

36. Collins KC, Kennedy NC, Clark A, Pomeroy VM. Kinematic Components of the Reach-to-Target Movement After Stroke for Focused Rehabilitation Interventions: Systematic Review and Meta-Analysis. Front Neurol. 2018;9. doi:10.3389/fneur.2018.00472

37. Bakhti KKA, Mottet D, Schweighofer N, Froger J, Laffont I. Proximal arm non-use when reaching after a stroke. Neurosci Lett. 2017;657:91–96. doi:10.1016/j.neulet.2017.07.055

38. Bakhti KKA, Laffont I, Muthalib M, Froger J, Mottet D. Kinect-based assessment of proximal arm non-use after a stroke. J Neuroeng Rehabil. 2018;15(1):104. doi:10.1186/s12984-018-0451-2

39. Faity G, Mottet D, Froger J. Validity and Reliability of Kinect v2 for Quantifying Upper Body Kinematics during Seated Reaching. Sensors. 2022;22(7):7. doi:10.3390/s22072735

40. Borg G a. V. Psychophysical bases of perceived exertion. Med Sci Sports Exerc. 1982;14(5):377–381.

41. de la Garanderie MP, Courtay A, Féral-Basin C, Rainville P, Gaveau J, Pageaux B. Perception of effort and the allocation of physical resources: A generalization to upper-limb motor tasks. Frontiers in Psychology. 2023;13. Accessed March 8, 2023. https://www.frontiersin.org/articles/10.3389/fpsyg.2022.974172

42. Selen LPJ, Beek PJ, van Dieën JH. Fatigue-induced changes of impedance and performance in target tracking. Exp Brain Res. 2007;181(1):99–108. doi:10.1007/s00221-007-0909-0

43. Fabbrini S, Casati G, Bonaiuti D. Is CIMT a rehabilitative practice for everyone? Predictive factors and feasibility. Eur J Phys Rehabil Med. 2014;50(5):10.

44. Tomita Y, Hasegawa S, Chida D, Asakura T, Usuda S. Association between self-perceived activity performance and upper limb functioning in subacute stroke. Physiother Res Int. 2022;27(3):e1946. doi:10.1002/pri.1946

45. de Rugy A, Loeb GE, Carroll TJ. Muscle Coordination Is Habitual Rather than Optimal. J Neurosci. 2012;32(21):7384–7391. doi:10.1523/JNEUROSCI.5792-11.2012

46. Ganesh G, Haruno M, Kawato M, Burdet E. Motor Memory and Local Minimization of Error and Effort, Not Global Optimization, Determine Motor Behavior. J Neurophysiol. 2010;104(1):382–390. doi:10.1152/jn.01058.2009

47. Michaelsen SM, Dannenbaum R, Levin MF. Task-Specific Training With Trunk Restraint on Arm Recovery in Stroke: Randomized Control Trial. Stroke. 2006;37(1):186–192. doi:10.1161/01.STR.0000196940.20446.c9

48. Kitago T, Liang J, Huang VS, et al. Improvement After Constraint-Induced Movement Therapy: Recovery of Normal Motor Control or Task-Specific Compensation? Neurorehabil Neural Repair. 2013;27(2):99–109. doi:10.1177/1545968312452631

49. Massie C, Malcolm MP, Greene D, Thaut M. The Effects of Constraint-Induced Therapy on Kinematic Outcomes and Compensatory Movement Patterns: An Exploratory Study. Arch Phys Med Rehabil. 2009;90(4):571–579. doi:10.1016/j.apmr.2008.09.574

50. Ellis MD, Sukal-Moulton T, Dewald JPA. Progressive Shoulder Abduction Loading is a Crucial Element of Arm Rehabilitation in Chronic Stroke. Neurorehabil Neural Repair. 2009;23(8):862–869. doi:10.1177/1545968309332927

