## Supplemental material for "Target Precision Shapes Trunk Compensation After Stroke: Neuromechanical Consequences for Upper Limb Rehabilitation"

Short title: Target Precision Shapes Trunk Compensation

### *SUPPLEMENTAL MATERIAL*

Supplemental Table S1. Repeated-measures ANOVAs on effects involving target size. Main effects of arm weight and arm-use instruction on trunk compensation, deltoid activation and perceived exertion were reported previously[S1]. In controls, target effects on movement time at each arm weight were tested with separate target × arm use ANOVAs.

| **Outcome / effect** | **Stroke (n=14), F(1,13)** | **p** | **η²p** | **Controls (n=19), F(1,18)** | **p** | **η²p** |
|---|---|---|---|---|---|---|
| **Trunk compensation** | | | | | | |
| Target | 8.166 | .013 | .39 | 0.057 | .814 | .003 |
| Arm weight × target | 0.084 | .777 | .01 | 0.681 | .420 | .04 |
| Target × arm use | 0.785 | .392 | .06 | 0.149 | .704 | .01 |
| Arm weight × target × arm use | 1.157 | .302 | .08 | 1.194 | .289 | .06 |
| **Anterior deltoid activation** | | | | | | |
| Target | 0.149 | .706 | .01 | 0.003 | .960 | <.001 |
| Arm weight × target | 0.610 | .449 | .04 | 0.255 | .620 | .01 |
| Target × arm use | 0.118 | .736 | .01 | 0.195 | .664 | .01 |
| Arm weight × target × arm use | 0.102 | .754 | .01 | 0.780 | .389 | .04 |
| **Movement time** | | | | | | |
| Target | 8.533 | .012 | .40 | 7.820 | .012 | .30 |
| Arm weight × target | 0.316 | .584 | .02 | 2.943 | .103 | .14 |
| Arm use | 0.006 | .939 | <.01 | 0.222 | .643 | .01 |
| Target × arm use | 0.012 | .913 | <.01 | 0.719 | .408 | .04 |
| Arm weight × target × arm use | 0.036 | .852 | <.01 | 3.909 | .064 | .18 |
| Target, normal arm weight (controls) | — | | | 0.158 | .695 | .01 |
| Target, weighted arm (controls) | — | | | 14.9 | .001 | .45 |
| **Perceived exertion** | | | | | | |
| Target | 4.304 | .058 | .25 | 9.500 | .006 | .35 |
| Arm weight × target | 4.304 | .058 | .25 | 5.444 | .031 | .23 |
| Target × arm use | 2.212 | .161 | .15 | 0.959 | .340 | .06 |
| Arm weight × target × arm use | 2.212 | .161 | .15 | 2.821 | .110 | .14 |

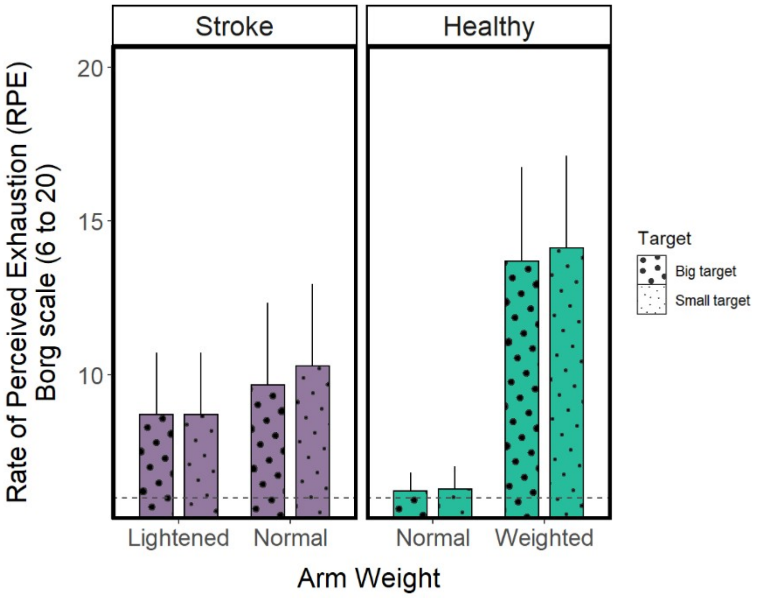


Supplemental Figure S1. Influence of target size and arm-weight context on perceived exertion. Perceived exertion was significantly larger for the small target in controls and showed a similar but non-significant trend in participants with stroke. Data are presented as mean (± standard deviation).

### Supplemental Reference

S1. Faity G, Mottet D, Froger J, Delorme M. Neuromechanics of Nonuse and Compensation: Implications for Rehabilitation After Stroke. *Neurorehabil Neural Repair*. 2026;40(8):615-630. doi:10.1177/15459683261445442
